# Nonlinear relationship between ionized calcium and 28-day mortality in patients with sepsis: A retrospective cohort study from MIMIC-IV database

**DOI:** 10.1101/2024.02.08.24302495

**Authors:** Zhanyao Liang, Yunting Chen, Yuanshen Zhou, Congqi Hu, Lu Chen, Fangfang Zhu

## Abstract

**Background:** This study aimed to investigate the linear and nonlinear relationships between ionized calcium levels and 28-day mortality in patients with sepsis in the intensive care unit (ICU) and to provide clinicians with a direction for laboratory index testing and a basis for a calcium supplementation program.

**Methods:** The data of patients with sepsis were extracted from the Medical Information Mart for Intensive Care (MIMIC-IV) database, iCa served as the exposure variable, and 28-day mortality served as the outcome variable. The relationship between iCa and 28-day mortality was investigated using multivariate binary logistic regression models, with adjustment for covariates. A generalized additive model (GAM) and smoothed curve fitting were used to investigate the non-linear relationship between iCa and 28-day mortality. A two-piecewise linear model was used to calculate the OR and 95% CI on either side of the inflection point.

**Results:** Patients with sepsis (n = 20,417) were included in the analysis, with an overall 28-day mortality rate of 19.73%. Sepsis patients were grouped into quartiles (Q1-Q4) according to ionized calcium levels, and after complete adjustment, the short-term mortality rate of sepsis patients in Q2–Q4 was significantly lower than that in the Q1 group (OR:0.78, 95% CI: 0.70–0.87. OR: 0.74, 95% CI: 0.66–0.83. OR: 0.81, 95% CI: 0.72–0.9. iCa and 28-day mortality displayed a U-shaped relationship among patients with sepsis. When iCa levels were less than 1.14 mmol/L, each unit increase corresponded to a 79% reduction in the risk of 28-day sepsis mortality(OR: 0.21, 95% CI: 0.13-0.35). Conversely, when iCa exceeded 1.14 mmol/L, each unit increase was linked to an 85% increase in the 28-day mortality risk (OR: 1.85, 95% CI: 1.08-3.16).

**Conclusion:** The relationship between iCa and 28-day mortality in patients with sepsis showed a u-shaped curve with an inflection point of 1.14 mmol/L, which is in the range of mild hypocalcemia.

## 1. Introduction

Sepsis, a life-threatening organ dysfunction caused by a dysregulated host response to infection ^[1]^, accounts for approximately 19.7% of all global deaths ^[2]^. Studies estimate that there are approximately 31.5 million new cases of sepsis and 5.3 million deaths worldwide each year, making it a major global public health problem ^[3]^.

Despite extensive efforts to discover new treatments and understand the underlying causes of sepsis, the mortality rate associated with this condition has not decreased ^[4,5]^. One challenge is the lack of reliable predictors for sepsis, which hinders accurate prediction of prognosis and leads to inadequate statistical power in clinical trials for new treatments. Therefore, it is crucial to explore simple and effective biomarkers to determine the prognosis of sepsis.

Abnormal serum calcium levels may be a marker of disease severity in critical illness ^[6–8]^. Hypocalcemia has been frequently observed in sepsis syndrome ^[9]^, and several studies have reported a statistically significant negative correlation between serum calcium levels and mortality rates in patients diagnosed with sepsis ^[10–12]^. However, other studies have reported that only extreme outliers of calcium levels are associated with sepsis mortality ^[13,14]^, so what is the real role of calcium in sepsis?

We hypothesize that inconsistencies in the findings of the aforementioned studies may be due to the following: (1) Some of the exposure factors in the studies were total calcium and some were ionized calcium (iCa). Approximately 50% of the total calcium is iCa, which is biologically active and less susceptible to other factors (e.g., albumin levels); therefore, iCa is preferred for assessment if available (2). Small sample sizes and inadequate adjustment for confounding factors resulted in less convincing conclusions. Therefore, this study used the extensive US multicenter sepsis database, Medical Information Mart for Intensive Care (MIMIC-IV), to investigate the association between iCa and the 28-day mortality risk in sepsis. The substantial sample size and extensive database information provided valuable statistical power, ensuring more robust and dependable findings that enhance our understanding of the relationship between iCa and the 28-day mortality risk in sepsis.

## 2. Methods

### 2.1 Study population

This study investigated health-related data obtained from the Medical Information Marketplace for Intensive Care IV (MIMIC-IV) database, which consists of extensive, high-quality medical records of patients in the Beth Israel Deaconess Medical Center (BIDMC), and was developed and managed by the MIT Computational Physiology Laboratory ^[15]^. The database is available for free download upon completion of an accredited course on its official website. Author (Lu Chen (RECORD 35931520) obtained access to the database and was responsible for data extraction. This study complied with the reports of studies conducted using the observation routine collected health data (RECORD) statement.

In this study, patients with sepsis attending BIDMC during 2008–2019 were extracted from the MIMIC-IV database based on ICD-9 codes recorded in the database to study the association between iCa levels and 28-day mortality in patients with sepsis. Patients with missing information on exposure variables were not included in this study.

Based on our clinical experience and reports in the literature, potential variables that were also analyzed included: demographic factors: gender (male/female), age at admission (years), ethnicity, charlson comorbidity index, SOFA score, heart rate, respiratory rate, and temperature on admission, use of mechanical ventilation, use of dialysis, use of norepinephrine, use of glucocorticosteroids (dexamethasone, methylprednisolone, hydrocortisone), use of vasoactive drugs (dopamine, dobutamine), immunoglobulins, antibiotics (carbapenem, cephalosporins, cillin-based drug, vancomycin).

### 2.2 Statistical analysis

Continuous variables are expressed as mean ± standard deviation (Gaussian distribution) or median (minimum, maximum) (skewed distribution), and categorical variables are expressed as rates. Because this was a cohort study, we divided the patients into 4 equal groups according to their iCa levels and observed the distribution of their baseline data among the different groups. One-way ANOVA (Gaussian distribution), Kruscal-WallisH (skewed distribution) test, and chi-square test (categorical variables) were used to determine any statistical differences between the means and proportions of the groups.

We used univariate and multivariate binary logistic regression models to observe the association between iCa levels and 28-day mortality in patients with sepsis. We used three models with a gradual degree of adjustment, including the unadjusted model (no covariates were adjusted, Model 1), the Minimally-adjusted model (adjusted only for demographic factors, Model 2), the Fully-adjusted model (adjusted for all the covariates presented in Table 1, Model 3) and present the Odds ratio value (OR) and 95% confidence intervals are presented.

**Table 1.**
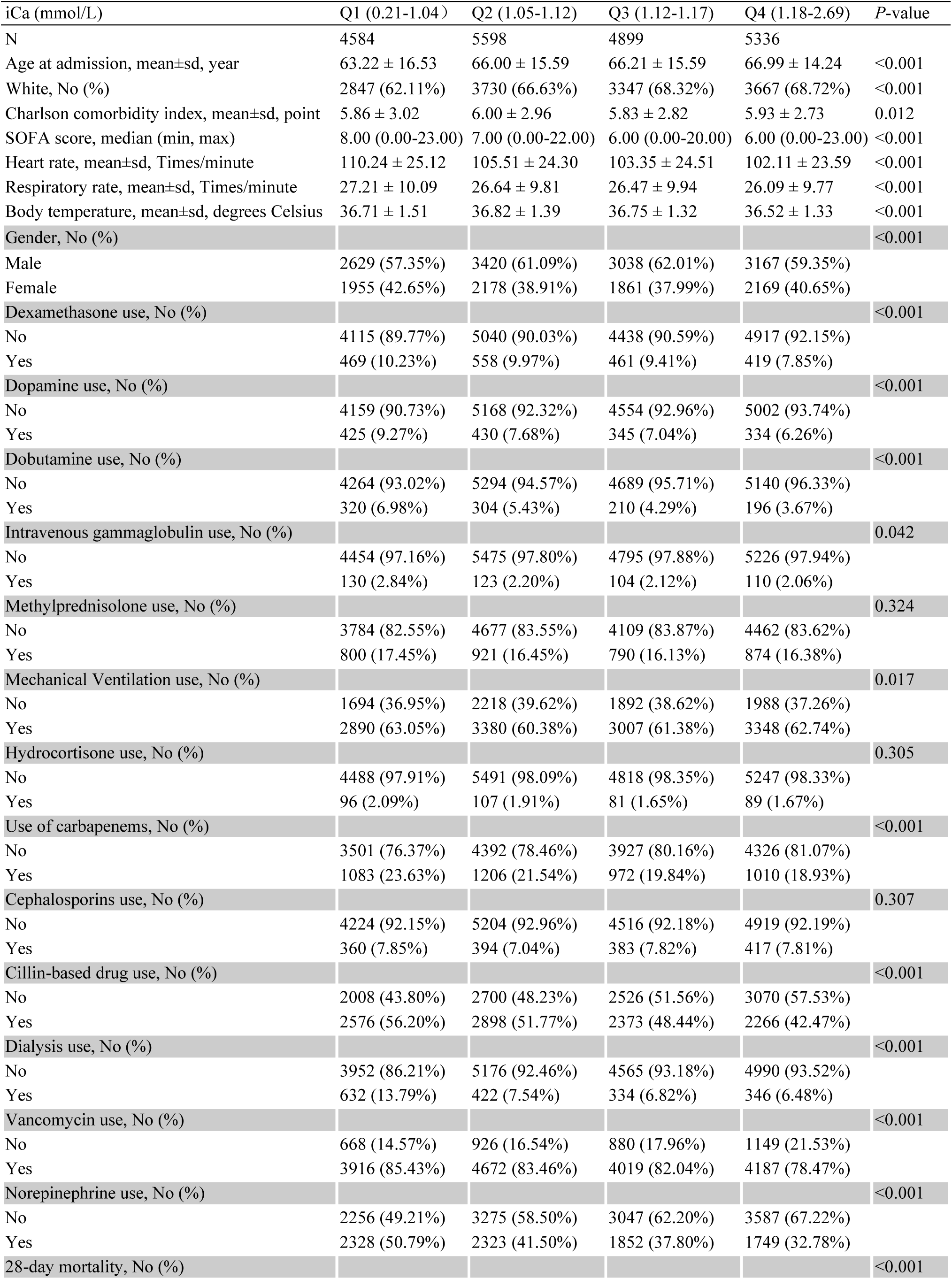
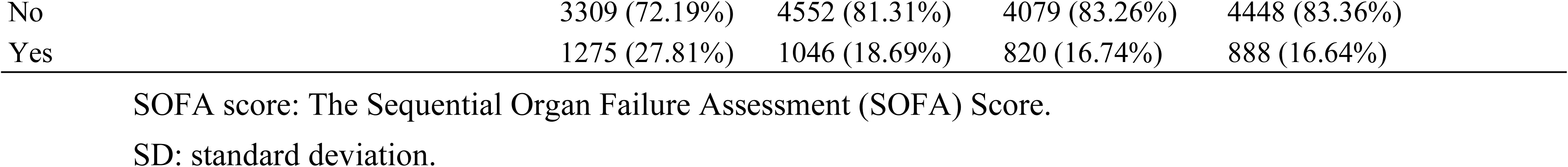
Description of baseline characteristics of patients with sepsis.

| iCa (mmol/L) | Q1 (0.21-1.04) | Q2 (1.05-1.12) | Q3 (1.12-1.17) | Q4 (1.18-2.69) | P-value |
| --- | --- | --- | --- | --- | --- |
| N | 4584 | 5598 | 4899 | 5336 |  |
| Age at admission, mean±sd, year | 63.22 ± 16.53 | 66.00 ± 15.59 | 66.21 ± 15.59 | 66.99 ± 14.24 | <0.001 |
| White, No (%) | 2847 (62.11%) | 3730 (66.63%) | 3347 (68.32%) | 3667 (68.72%) | <0.001 |
| Charlson comorbidity index, mean±sd, point | 5.86 ± 3.02 | 6.00 ± 2.96 | 5.83 ± 2.82 | 5.93 ± 2.73 | 0.012 |
| SOFA score, median (min, max) | 8.00 (0.00-23.00) | 7.00 (0.00-22.00) | 6.00 (0.00-20.00) | 6.00 (0.00-23.00) | <0.001 |
| Heart rate, mean±sd, Times/minute | 110.24 ± 25.12 | 105.51 ± 24.30 | 103.35 ± 24.51 | 102.11 ± 23.59 | <0.001 |
| Respiratory rate, mean±sd, Times/minute | 27.21 ± 10.09 | 26.64 ± 9.81 | 26.47 ± 9.94 | 26.09 ± 9.77 | <0.001 |
| Body temperature, mean±sd, degrees Celsius | 36.71 ± 1.51 | 36.82 ± 1.39 | 36.75 ± 1.32 | 36.52 ± 1.33 | <0.001 |
| Gender, No (%) |  |  |  |  | <0.001 |
| Male | 2629 (57.35%) | 3420 (61.09%) | 3038 (62.01%) | 3167 (59.35%) |  |
| Female | 1955 (42.65%) | 2178 (38.91%) | 1861 (37.99%) | 2169 (40.65%) |  |
| Dexamethasone use, No (%) |  |  |  |  | <0.001 |
| No | 4115 (89.77%) | 5040 (90.03%) | 4438 (90.59%) | 4917 (92.15%) |  |
| Yes | 469 (10.23%) | 558 (9.97%) | 461 (9.41%) | 419 (7.85%) |  |
| Dopamine use, No (%) |  |  |  |  | <0.001 |
| No | 4159 (90.73%) | 5168 (92.32%) | 4554 (92.96%) | 5002 (93.74%) |  |
| Yes | 425 (9.27%) | 430 (7.68%) | 345 (7.04%) | 334 (6.26%) |  |
| Dobutamine use, No (%) |  |  |  |  | <0.001 |
| No | 4264 (93.02%) | 5294 (94.57%) | 4689 (95.71%) | 5140 (96.33%) |  |
| Yes | 320 (6.98%) | 304 (5.43%) | 210 (4.29%) | 196 (3.67%) |  |
| Intravenous gammaglobulin use, No (%) |  |  |  |  | 0.042 |
| No | 4454 (97.16%) | 5475 (97.80%) | 4795 (97.88%) | 5226 (97.94%) |  |
| Yes | 130 (2.84%) | 123 (2.20%) | 104 (2.12%) | 110 (2.06%) |  |
| Methylprednisolone use, No (%) |  |  |  |  | 0.324 |
| No | 3784 (82.55%) | 4677 (83.55%) | 4109 (83.87%) | 4462 (83.62%) |  |
| Yes | 800 (17.45%) | 921 (16.45%) | 790 (16.13%) | 874 (16.38%) |  |
| Mechanical Ventilation use, No (%) |  |  |  |  | 0.017 |
| No | 1694 (36.95%) | 2218 (39.62%) | 1892 (38.62%) | 1988 (37.26%) |  |
| Yes | 2890 (63.05%) | 3380 (60.38%) | 3007 (61.38%) | 3348 (62.74%) |  |
| Hydrocortisone use, No (%) |  |  |  |  | 0.305 |
| No | 4488 (97.91%) | 5491 (98.09%) | 4818 (98.35%) | 5247 (98.33%) |  |
| Yes | 96 (2.09%) | 107 (1.91%) | 81 (1.65%) | 89 (1.67%) |  |
| Use of carbapenems, No (%) |  |  |  |  | <0.001 |
| No | 3501 (76.37%) | 4392 (78.46%) | 3927 (80.16%) | 4326 (81.07%) |  |
| Yes | 1083 (23.63%) | 1206 (21.54%) | 972 (19.84%) | 1010 (18.93%) |  |
| Cephalosporins use, No (%) |  |  |  |  | 0.307 |
| No | 4224 (92.15%) | 5204 (92.96%) | 4516 (92.18%) | 4919 (92.19%) |  |
| Yes | 360 (7.85%) | 394 (7.04%) | 383 (7.82%) | 417 (7.81%) |  |
| Cillin-based drug use, No (%) |  |  |  |  | <0.001 |
| No | 2008 (43.80%) | 2700 (48.23%) | 2526 (51.56%) | 3070 (57.53%) |  |
| Yes | 2576 (56.20%) | 2898 (51.77%) | 2373 (48.44%) | 2266 (42.47%) |  |
| Dialysis use, No (%) |  |  |  |  | <0.001 |
| No | 3952 (86.21%) | 5176 (92.46%) | 4565 (93.18%) | 4990 (93.52%) |  |
| Yes | 632 (13.79%) | 422 (7.54%) | 334 (6.82%) | 346 (6.48%) |  |
| Vancomycin use, No (%) |  |  |  |  | <0.001 |
| No | 668 (14.57%) | 926 (16.54%) | 880 (17.96%) | 1149 (21.53%) |  |
| Yes | 3916 (85.43%) | 4672 (83.46%) | 4019 (82.04%) | 4187 (78.47%) |  |
| Norepinephrine use, No (%) |  |  |  |  | <0.001 |
| No | 2256 (49.21%) | 3275 (58.50%) | 3047 (62.20%) | 3587 (67.22%) |  |
| Yes | 2328 (50.79%) | 2323 (41.50%) | 1852 (37.80%) | 1749 (32.78%) |  |
| 28-day mortality, No (%) |  |  |  |  | <0.001 |

|  |  |  |  |  |
| --- | --- | --- | --- | --- |
| No | 3309 (72.19%) | 4552 (81.31%) | 4079 (83.26%) | 4448 (83.36%) |
| Yes | 1275 (27.81%) | 1046 (18.69%) | 820 (16.74%) | 888 (16.64%) |

SOFA score: The Sequential Organ Failure Assessment (SOFA) Score.
SD: standard deviation.

In addition, for the sensitivity analysis, we transformed calcium ion levels from a continuous variable to a categorical variable (quartile) and calculated *P* for trend, with the aim of observing whether the results were robust when calcium ion levels were used as a continuous variable versus as a categorical variable.

Because iCa levels are continuous variables, the possibility of a nonlinear association cannot be excluded. Given the incompetence of the binary logistic regression model in dealing with nonlinear associations, we used a generalized additive model (GAM) and smoothed curve fitting to observe the association between ionic calcium levels and 28-day mortality in sepsis. If a nonlinear association existed, we first calculated the inflection point values using a recursive algorithm, and then calculated OR values and 95% confidence intervals on both sides of the inflection point using a two-piecewise linear model.

All analyses were performed using the statistical packages R (http://www.R-project.org, The R Foundation) and EmpowerStats (http://www. mpowerstats.com, X&Y Solutions, Inc, Boston, MA). *P* values < 0.05 (two-sided) were considered statistically significant.

## 3. Results

### 3.1 Screening process

In this study, we enrolled 377,207 patients from the MIMIC-IV database. Among these, 342,197 were non-septic patients, while 14,593 had missing information on iCa. Consequently, 20,417 cases were considered for the final analysis. The flow chart of patient selection is depicted in Fig .1.

**Fig. 1.**
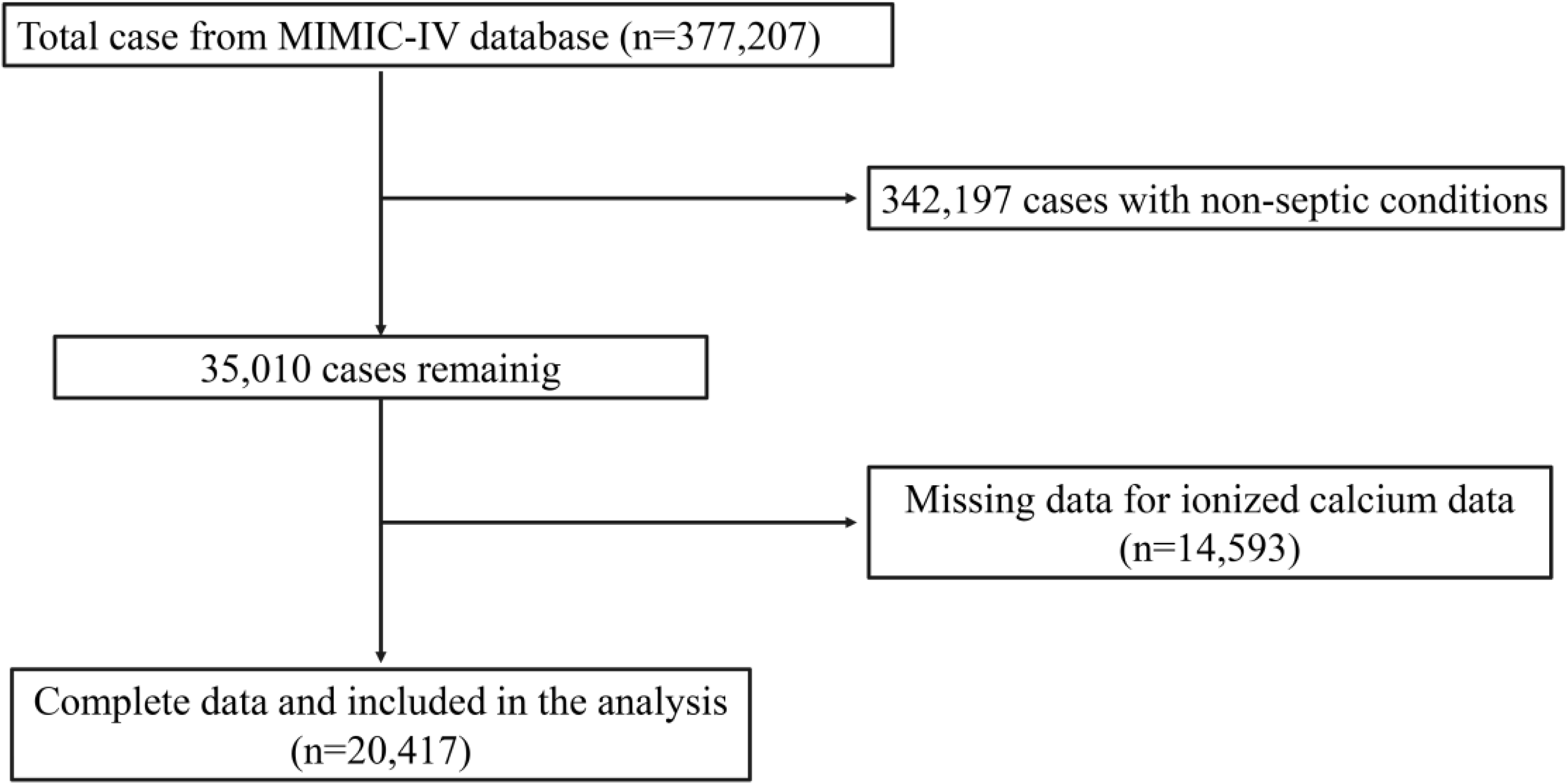
Flow chart of patient selection.

### 3.2 Baseline characteristics

Patient baseline characteristics are summarized in Table 1. iCa levels in the entire population were segmented into four groups (Q1 to Q4) based on quartile divisions. Trends in the distribution of each variable across different subgroups were observed following this segmentation. Analysis revealed that, relative to the Q1 group, patients in the Q2, Q3, and Q4 groups were typically older, had a higher representation of white ethnicity, a higher Charlson Comorbidity Index, and more frequent use of dexamethasone and dopamine. These groups also exhibited lower SOFA scores, heart rate, and respiratory rate. They received a lower percentage of treatments such as methylprednisolone, hydrocortisone, norepinephrine, intravenous gammaglobulin, cephalosporins, cillin-based drugs, carbapenems, vancomycin, mechanical ventilation, and dialysis and demonstrated a lower 28-day mortality rate. The 28-day mortality rate for patients with sepsis was 19.73%.

### 3.3 Relationship between iCa and 28-day mortality in patients with sepsis using non-adjusted and adjusted models

Various covariate adjustment strategies were employed to elucidate the relationship between iCa and 28-day mortality in patients with sepsis. The non-adjusted and adjusted models are detailed in Table 2. The non-adjusted model revealed that for each unit increase in iCa, the 28-day mortality risk in patients with sepsis was reduced by 86% (OR: 0.14, 95%CI: 0.11-0.19, *P*<0.001). After adjusting for sex, age at admission, and ethnicity in Model 1, the trend remained consistent (OR: 0.13, 95%CI: 0.09-0.17, *P* <0.001). In Model 2, which was adjusted for all covariates listed in Table 1, the 28-day mortality risk was reduced by 41% (OR: 0.59, 95%CI: 0.43-0.82, *P* <0.001). Sensitivity analysis was also conducted with iCa treated as a categorical variable (quartile), and a similar trend was observed (*P* for trend <0.0001).

**Table 2.** Results of univariate and multivariable logistic regression analysis.

| Exposure | Model 0<br>OR, 95%CI, *P* value | Model 1<br>OR, 95%CI, *P* value | Model 2<br>OR, 95%CI, *P* value | GAM<br>OR, 95%CI, *P* value |
| --- | --- | --- | --- | --- |
| iCa (mmol/L) | 0.14 (0.11, 0.19) <0.0001 | 0.13 (0.09, 0.17) <0.0001 | 0.59 (0.43, 0.82) 0.0014 | 0.64 (0.46, 0.88) 0.0064 |
| iCa quartile |  |  |  |  |
| Q1 | reference | reference | reference | reference |
| Q2 | 0.60 (0.54, 0.65) <0.0001 | 0.57 (0.52, 0.63) <0.0001 | 0.78 (0.70, 0.87) <0.0001 | 0.79 (0.71, 0.88) <0.0001 |
| Q3 | 0.52 (0.47, 0.58) <0.0001 | 0.50 (0.45, 0.55) <0.0001 | 0.74 (0.66, 0.83) <0.0001 | 0.75 (0.67, 0.84) <0.0001 |
| Q4 | 0.52 (0.47, 0.57) <0.0001 | 0.50 (0.45, 0.55) <0.0001 | 0.81 (0.72, 0.90) 0.0002 | 0.83 (0.74, 0.93) 0.0017 |
| *P* for trend | <0.0001 | <0.0001 | <0.0001 | 0.0009 |

OR: Odds ratio.
95% CI: 95% confidence interval.
Model 0: no covariates were adjusted for.
Model 1: We adjusted only for sex, age at admission, and ethnicity.
Model 2: We adjusted for all covariates presented in Table 1.
GAM: All variables listed in Model II were adjusted. However, continuous variables were adjusted as non-linearity.

### 3.4 Non-linear relationship between iCa and 28-day mortality

A generalized additive model and smoothed curve fitting were used to examine the nonlinear relationship between iCa and 28-day mortality. The findings suggested a U-shaped association between iCa and 28-day mortality after adjusting for covariates (The adjustment strategy aligns with model2 in Table 2) (Refer to Fig .2). The inflection point for iCa was calculated to be 1.14 mmol/L using a two-piecewise linear model and a recursive algorithm. For iCa levels less than 1.14 mmol/L, each unit increase corresponded to a 79% reduction in the risk of 28-day sepsis mortality (OR: 0.21, 95% CI: 0.13 to 0.35, *P* < 0.0001). Conversely, when iCa exceeded 1.14 mmol/L, each unit increase was linked to an 85% rise in the 28-day mortality risk (OR: 1.85, 95% CI: 1.08 to 3.16, *P* < 0.05) (Table 3).

**Fig. 2.**
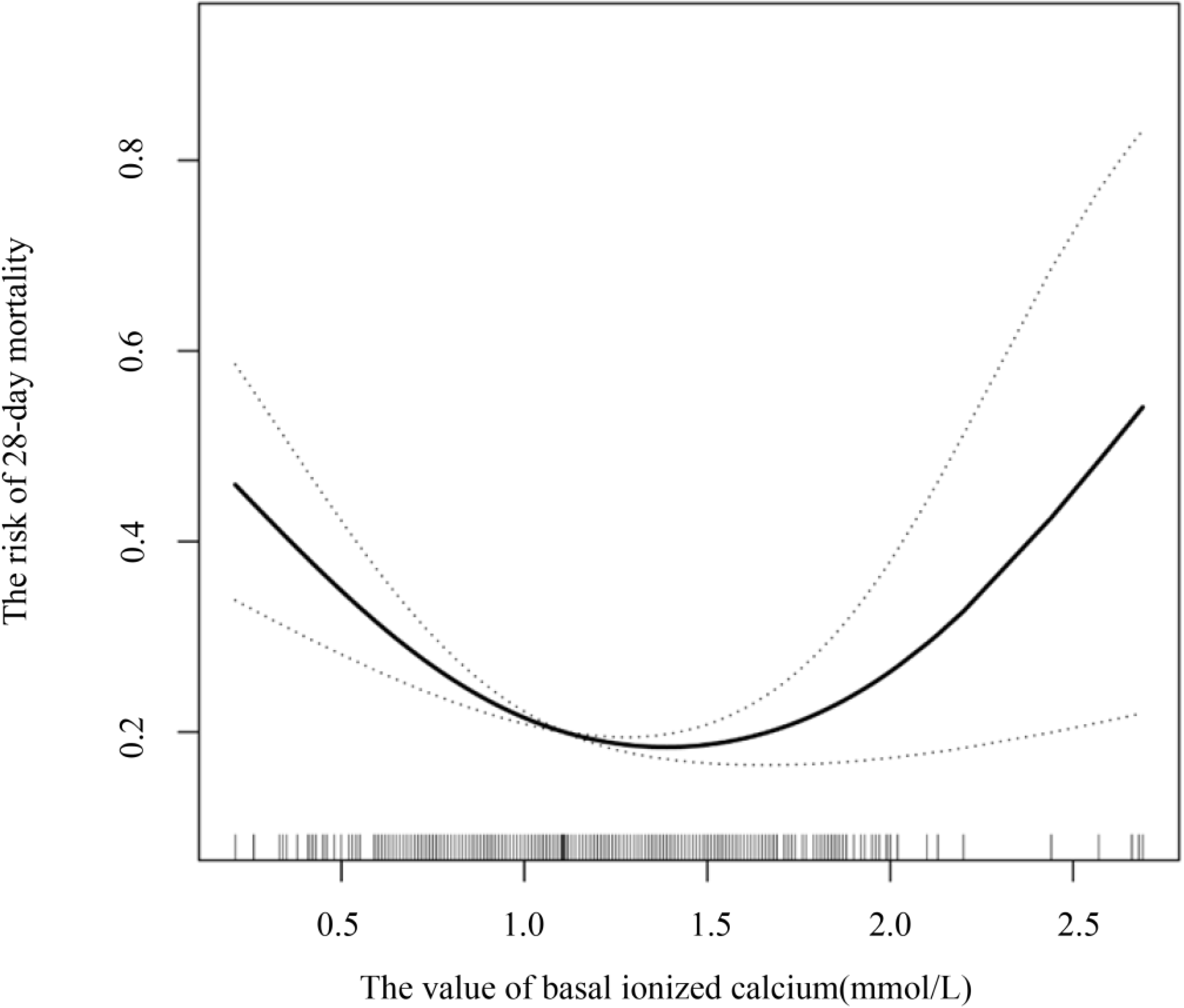
Correlation between iCa levels and 28-day mortality probability in sepsis patients. The solid line represents the smoothed curve, while the dashed line indicates the 95% confidence interval.

**Table 3.** Explanation of the non-linear association between iCa and the risk of 28-day mortality in sepsis patient.

| Outcome: | OR, 95%CI, *P* value |
| --- | --- |
| Fitting model using standard logistic regression | 0.59 (0.43, 0.82) 0.0014 |
| Fitting model using two-piecewise linear model |  |
| Inflection point | 1.14 (1.12 to 1.15) |
| ≤ inflection point | 0.21 (0.13, 0.35) <0.0001 |
| > inflection point | 1.85 (1.08, 3.16) 0.0247 |
| Log-likelihood ration test | <0.001 |

The adjustment strategy is the same as model 2 in Table 2.

## 4. Discussion

In the present study, we evaluated the association between iCa levels and 28-day mortality in patients with sepsis from the MIMIC IV database, which, to the best of our knowledge, is the largest sample size study on iCa and short-term mortality in sepsis. The results of this study indicate that iCa levels are independently associated with 28-day mortality in patients with sepsis. They remained strongly correlated even after correction for confounding risk factors. Importantly, by GAM modeling and smoothed curve fitting, we found a U-shaped relationship between iCa levels and 28-day mortality in patients with sepsis, i.e., when iCa levels were less than 1.14 mmol/L, each unit increase corresponded to a 79% reduction in the risk of 28-day sepsis mortality. Conversely, when iCa exceeded 1.14 mmol/L, each unit increase was linked to an 85% increase in the 28-day mortality risk.

The rapid progression and high mortality of sepsis implies the necessity of identifying prognostic indicators associated with its outcome. It is recommended that scholars further explore serum markers and prognostic correlates to help clinicians intervene early ^[16,17]^. Calcium is an essential trace element in the human body that is responsible for physiological functions including cell signaling, neurotransmission, and muscle contraction ^[6,18,19]^. Calcium disorders are uncommon in the general population, except in special cases (e.g., renal failure), and are therefore often neglected ^[20,21]^. Nevertheless, calcium derangements in sepsis are prevalent and should be taken seriously ^[22,23]^. Total calcium levels are affected by a variety of factors, such as hypoalbuminemia or hypoalbuminemia, and may be less accurate (in the presence of renal disease, acid-base disturbances, or anionic perturbations), even after adjustment by algorithms ^[24–26]^. In these cases, iCa measurement may be preferred if feasible and can improve more accurate judgment for the clinician; however, there is a paucity of high-quality studies on iCa and sepsis.

In this study, we found that iCa was negatively associated with 28-day mortality from sepsis when it less than 1.14 mmol/L. A retrospective study on elderly patients with sepsis showed that the serum calcium levels of septic patients were lower than those of the control group and that the severity of sepsis was associated with lower serum calcium levels ^[11]^. A retrospective study on pediatric sepsis also revealed that among children with sepsis, those with low blood calcium levels had a higher incidence of organ dysfunction than those with normal blood calcium levels ^[11]^. These studies increase our confidence in our findings.

The normal range of iCa in adults is 1.20–1.40 mmoL/L (4.8-5.6 mg/dL) ^[6]^. In this study, we found that iCa was below the normal range in most septicemic patients. Furthermore, sepsis mortality was lowest at an iCa level of 1.14 mmol/L, which is slightly below the normal reference range for iCa. This suggests that mild hypocalcemia may be protective against sepsis. A retrospective study ^[27]^ found that mild hypocalcemia may have a protective effect in septic patients, and calcium supplementation may have both positive and negative effects on mortality depending on the severity of the disease. Aberegg also suggested that low iCa levels may be protective in critically ill patients, whereas attempts to correct low levels may be harmful [6]. Our findings are partially consistent with the perspectives of the aforementioned researchers. In a sepsis animal model, calcium supplementation was associated with decreased survival, whereas no deaths were observed in hypocalcemic animals ^[28]^. Whether low iCa is a protective mechanism or a consequence of metabolic imbalance in sepsis requires further mechanistic studies. Because of the complex and wide-ranging effects of iCa, this result should be interpreted cautiously.

Severe hypercalcemia is associated with an increased risk of death, which is attributable in most cases to primary hyperparathyroidism or malignancy ^[29–31]^. However, by exploring the nonlinear relationship between iCa and sepsis, we found a significant positive correlation between elevated iCa levels and short-term mortality in patients with sepsis once iCa was greater than 1.14 mmol/L, even if severe hypercalcemia levels were not reached ( iCa < 2.5 mmol/L). The mechanism, although not fully delineated, may be due to several factors, including cardiovascular risk. Elevated iCa can lead to increased cardiac load, causing arrhythmias, elevated blood pressure, or increased myocardial contractility, which can further disrupt cardiovascular function in patients with sepsis ^[32–34]^. (2) Thrombosis formation: Elevated calcium levels enhance coagulation factor activity and increase blood viscosity, thereby increasing the risk of thrombosis ^[35–37]^. (3) Immune dysregulation: Hypercalcemia impairs immune cell function and disrupts immune system regulation, which increases the severity of infection, exacerbates sepsis, and increases the risk of _death_ ^[38,39]^.

Our study has some advantages. First, we used advanced algorithms such as generalized additive modeling (GAM), smoothed curve fitting, and two-segment linear models, which allow for a more accurate determination of the curvilinear and quantitative relationship between iCa and the 28-day risk of death in patients with sepsis. Second, we adjusted for various factors related to short-term prognosis, including demographic information (such as gender, age, race, etc.), Charlson comorbidity index, SOFA score, medication use related to sepsis prognosis (such as steroids, vasopressor antibiotics, etc.), and invasive procedures. This adjustment helps eliminate interference from other factors and makes the results more reliable. Third, we chose to study physiologically active iCa rather than total serum calcium. This is because when patients have fluid overload, chronic diseases, or malnutrition, the total calcium in the plasma may decrease, but the iCa level remains normal. Studying iCa allows for a more accurate assessment of calcium concentration and its correlation with short-term prognosis.

However, some limitations should also be discussed. First, our study primarily focused on the evaluation of iCa levels at the time of initial admission to the intensive care unit (ICU), recognizing the potential limitations associated with this single measurement. Nonetheless, we discovered a substantial association between iCa levels and mortality among septic patients. Second, because the data come mainly from patients in the United States, more clinical analysis is needed to test whether our results are applicable to patients with sepsis in other countries. Third, as an observational study, we can only establish associations and cannot determine a causal relationship between iCa and the risk of death from sepsis. Hence, this study should serve as the basis for future well-designed studies to evaluate the impact of iCa counts on mortality and causality.

## 5. Conclusion

The relationship between iCa and 28-day mortality in patients with sepsis showed a u-shaped curve with an inflection point of 1.14 mmol/L, which is in the range of mild hypocalcemia. When iCa levels were 1.14 mmol/L, each unit increase corresponded to a 79% reduction in the risk of 28-day sepsis mortality. Conversely, when iCa exceeded 1.14 mmol/L, each unit increase was linked to an 85% increase in the 28-day mortality risk. These findings are potentially valuable in assessing the prognosis of patients with sepsis and in adjusting the testing and treatment program for patients with sepsis.

## Ethics statement

The data for this study were obtained from the MIMIC-IV public database on the Internet, which has been approved by the institutional review boards of Beth Israel Deaconess Medical Center in Boston, Massachusetts (2001-P-001699/14), and the Massachusetts Institute of Technology (0403000206). Informed consent was abrogated because the data were public and the patients’ personal information was uncertain.

## Author contributions

All authors read and approved the final manuscript. ZYS were responsible for the study concept and design. CL were responsibility for collecting the data, statistical analysis. HCQ were responsible for literature retrieval. LZY and CYT were responsible for drafting the manuscript. ZFF was responsible for critical reading of a final version of the manuscript.

## Publisher’s note

All claims expressed in this article are solely those of the authors and do not necessarily represent those of their affiliated organizations, or those of the publisher, the editors and the reviewers. Any product that may be evaluated in this article, or claim that may be made by its manufacturer, is not guaranteed or endorsed by the publisher.

## Data Availability

The data are available on the MIMIC-IV; database website at https://mimic-IV.mit.edu/.

https://mimic-&#8547;.mit.edu/.

## References

[1] C.W. Seymour, V.X. Liu, T.J. Iwashyna, et al., Assessment of clinical criteria for sepsis: for the third international consensus definitions for sepsis and septic shock (sepsis-3), JAMA-J. Am. Med. Assoc. 315 (8) (2016) 762–774, 10.1001/jama.2016.0288.

[2] K.E. Rudd, S.C. Johnson, K.M. Agesa, et al., Global, regional, and national sepsis incidence and mortality, 1990-2017: analysis for the global burden of disease study, Lancet 395 (10219) (2020) 200-211, 10.1016/S0140-6736(19)32989-7.

[3] M. van den Berg, F.E. van Beuningen, J.C. Ter Maaten, H.R. Bouma, Hospital-related costs of sepsis around the world: a systematic review exploring the economic burden of sepsis, J. Crit. Care 712022) 154096, 10.1016/j.jcrc.2022.154096.

[4] C. Chiu, M. Legrand, Epidemiology of sepsis and septic shock, Curr. Opin. Anesthesiol. 34 (2) (2021) 71–76, 10.1097/ACO.0000000000000958.

[5] R. Luhr, Y. Cao, B. Soderquist, S. Cajander, Trends in sepsis mortality over time in randomised sepsis trials: a systematic literature review and meta-analysis of mortality in the control arm, 2002-2016, Crit. Care 23 (1) (2019) 241, 10.1186/s13054-019-2528-0.

[6] S.K. Aberegg, Ionized calcium in the icu: should it be measured and corrected? Chest 149 (3) (2016) 846–855, 10.1016/j.chest.2015.12.001.

[7] T. Peng, X. Peng, M. Huang, et al., Serum calcium as an indicator of persistent organ failure in acute pancreatitis, Am. J. Emerg. Med. 35 (7) (2017) 978–982, 10.1016/j.ajem.2017.02.006.

[8] J.K. Sun, W.H. Zhang, L. Zou, et al., Serum calcium as a biomarker of clinical severity and prognosis in patients with coronavirus disease 2019, Aging (Albany NY) 12 (12) (2020) 11287–11295, 10.18632/aging.103526.

[9] J.R. Zivin, T. Gooley, R.A. Zager, M.J. Ryan, Hypocalcemia: a pervasive metabolic abnormality in the critically ill, Am. J. Kidney Dis. 37 (4) (2001) 689–698, 10.1016/s0272-6386(01)80116-5.

[10] H. Li, J. Chen, Y. Hu, X. Cai, D. Tang, P. Zhang, Clinical value of serum calcium in elderly patients with sepsis, Am. J. Emerg. Med. 522022) 208-211, 10.1016/j.ajem.2021.12.019.

[11] Y. Liu, Y. Chai, Z. Rong, Y. Chen, Prognostic value of ionized calcium levels in neonatal sepsis, Ann. Nutr. Metab. 76 (3) (2020) 193–200, 10.1159/000508685.

[12] X. Zheng, Y. Li, Q. Cheng, L. Wang, Predictive value of ionized calcium for prognosis of sepsis in very low birth weight infants, J. Inflamm. Res. 152022) 3749-3760, 10.2147/JIR.S369431.

[13] M. Egi, I. Kim, A. Nichol, et al., Ionized calcium concentration and outcome in critical illness, Crit. Care Med. 39 (2) (2011) 314–321, 10.1097/CCM.0b013e3181ffe23e.

[14] D. Yan, X. Xie, X. Fu, et al., U-shaped association between serum calcium levels and 28-day mortality in patients with sepsis: a retrospective analysis of the mimic-iii database, Shock 60 (4) (2023) 525–533, 10.1097/SHK.0000000000002203.

[15] A. Johnson, L. Bulgarelli, L. Shen, et al., Mimic-iv, a freely accessible electronic health record dataset, Sci. Data 10 (1) (2023) 1, 10.1038/s41597-022-01899-x.

[16] G. Barber, J. Tanic, A. Leligdowicz, Circulating protein and lipid markers of early sepsis diagnosis and prognosis: a scoping review, Curr. Opin. Lipidology 34 (2) (2023) 70–81, 10.1097/MOL.0000000000000870.

[17] A.C. Vazquez, L. Arriaga-Pizano, E. Ferat-Osorio, Cellular markers of immunosuppression in sepsis, Arch. Med. Res. 52 (8) (2021) 828–835, 10.1016/j.arcmed.2021.10.001.

[18] L. Bai, S. Sun, Y. Sun, F. Wang, A. Nishiyama, N-type calcium channel and renal injury, Int. Urol. Nephrol. 54 (11) (2022) 2871–2879, 10.1007/s11255-022-03183-8.

[19] P.D. Nguyen, I. Gooijers, G. Campostrini, et al., Interplay between calcium and sarcomeres directs cardiomyocyte maturation during regeneration, Science 380 (6646) (2023) 758-764, 10.1126/science.abo6718.

[20] A. Kelly, M.A. Levine, Hypocalcemia in the critically ill patient, J. Intensive Care Med. 28 (3) (2013) 166–177, 10.1177/0885066611411543.

[21] M.B. Gallo, G. Aghagoli, S.L. Hu, C.M. Massoud, L. Robinson-Bostom, Calciphylaxis and kidney disease: a review, Am. J. Kidney Dis. 81 (2) (2023) 232–239, 10.1053/j.ajkd.2022.06.011.

[22] A. Sood, G. Singh, T.G. Singh, K. Gupta, Pathological role of the calcium-sensing receptor in sepsis-induced hypotensive shock: therapeutic possibilities and unanswered questions, Drug Dev. Res. 83 (6) (2022) 1241–1245, 10.1002/ddr.21959.

[23] H. Yang, Z. Zhang, Sepsis-induced myocardial dysfunction: the role of mitochondrial dysfunction, Inflamm. Res. 70 (4) (2021) 379–387, 10.1007/s00011-021-01447-0.

[24] I.A. Lian, A. Asberg, Should total calcium be adjusted for albumin? A retrospective observational study of laboratory data from central norway, BMJ Open 8 (4) (2018) e017703, 10.1136/bmjopen-2017-017703.

[25] R.B. Payne, A.J. Little, R.B. Williams, J.R. Milner, Interpretation of serum calcium in patients with abnormal serum proteins, Br Med J 4 (5893) (1973) 643-646, 10.1136/bmj.4.5893.643.

[26] J.D. Smith, S. Wilson, H.G. Schneider, Misclassification of calcium status based on albumin-adjusted calcium: studies in a tertiary hospital setting, Clin. Chem. 64 (12) (2018) 1713–1722, 10.1373/clinchem.2018.291377.

[27] W. He, L. Huang, H. Luo, et al., The positive and negative effects of calcium supplementation on mortality in septic icu patients depend on disease severity: a retrospective study from the mimic-iii, Crit. Care Res. Pract. 20222022) 2520695, 10.1155/2022/2520695.

[28] D.S. Malcolm, G.P. Zaloga, J.W. Holaday, Calcium administration increases the mortality of endotoxic shock in rats, Crit. Care Med. 17 (9) (1989) 900–903, 10.1097/00003246-198909000-00012.

[29] T.P. Jacobs, J.P. Bilezikian, Clinical review: rare causes of hypercalcemia, J. Clin. Endocrinol. Metab. 90 (11) (2005) 6316–6322, 10.1210/jc.2005-0675.

[30] B. Wang, Y. Gong, B. Ying, B. Cheng, Association of initial serum total calcium concentration with mortality in critical illness, Biomed Res. Int. 20182018) 7648506, 10.1155/2018/7648506.

[31] M.I. Hu, Hypercalcemia of malignancy, Endocrinol. Metabol. Clin. North Amer. 50 (4) (2021) 721-728, 10.1016/j.ecl.2021.07.003.

[32] G. Bkaily, D. Jacques, Calcium homeostasis, transporters, and blockers in health and diseases of the cardiovascular system, Int. J. Mol. Sci. 24 (10) (2023), 10.3390/ijms24108803.

[33] C.J. Kobylecki, B.G. Nordestgaard, S. Afzal, Plasma ionized calcium and risk of cardiovascular disease: 106 774 individuals from the copenhagen general population study, Clin. Chem. 67 (1) (2021) 265–275, 10.1093/clinchem/hvaa245.

[34] B. Thompson, M. Waterhouse, D.R. English, et al., Vitamin d supplementation and major cardiovascular events: d-health randomised controlled trial, BMJ-British Medical Journal 3812023) e075230, 10.1136/bmj-2023-075230.

[35] T. Koufakis, V. Antonopoulou, M. Grammatiki, et al., The relationship between primary hyperparathyroidism and thrombotic events: report of three cases and a review of potential mechanisms, Int J Hematol Oncol Stem Cell Res 12 (3) (2018) 175–180.

[36] G. Yorulmaz, A.T. Kalkan, A. Akalin, et al., Effect of hyperparathyroidism on coagulation: a global assessment by modified rotation thromboelastogram (rotem), Turk. J. Med. Sci. 51 (6) (2021) 2897–2902, 10.3906/sag-2012-247.

[37] G. Khattar, F.S. Siddiqui, R. Grovu, et al., The calcium-clot connection: investigating the association between primary hyperparathyroidism and acute venous thromboembolism, J. Thromb. Thrombolysis 2023), 10.1007/s11239-023-02906-7.

[38] G.N. Hendy, L. Canaff, Calcium-sensing receptor, proinflammatory cytokines and calcium homeostasis, Semin. Cell Dev. Biol. 492016) 37-43, 10.1016/j.semcdb.2015.11.006.

[39] H. Izzedine, T. Chazal, R. Wanchoo, K.D. Jhaveri, Immune checkpoint inhibitor-associated hypercalcaemia, Nephrol. Dial. Transplant. 37 (9) (2022) 1598–1608, 10.1093/ndt/gfaa326.

