## Supplementary table for "Nonlinear relationship between ionized calcium and 28-day mortality in patients with sepsis: A retrospective cohort study from MIMIC-IV database"

Description of missing data.

| Variables | Missing | Without missing | missing rate |
| --- | --- | --- | --- |
| Age at admission | 35010 | 0 | 0 |
| Charlson comorbidity index | 35010 | 0 | 0 |
| SOFA score | 35010 | 0 | 0 |
| Cephalosporins use | 35010 | 0 | 0 |
| Cillin-based drug use | 35010 | 0 | 0 |
| 28-day mortality | 35010 | 0 | 0 |
| Dexamethasone use | 35010 | 0 | 0 |
| Dopamine use | 35010 | 0 | 0 |
| Dobutamine use | 35010 | 0 | 0 |
| ethnicity | 35010 | 0 | 0 |
| Gender | 35010 | 0 | 0 |
| Intravenous gammaglobulin use | 35010 | 0 | 0 |
| Methylprednisolone use | 35010 | 0 | 0 |
| Mechanical Ventilation use | 35010 | 0 | 0 |
| Hydrocortisone use | 35010 | 0 | 0 |
| Use of carbapenems | 35010 | 0 | 0 |
| Heart rate | 34983 | 27 | 0.000771803 |
| Respiratory rate | 34968 | 42 | 0.001201098 |
| Dialysis use | 35010 | 0 | 0 |
| Body temperature | 33568 | 442 | 0.013167302 |
| Vancomycin use | 35010 | 0 | 0 |
